# Attempt to replicate voxel-based morphometry analysis in fibromyalgia: Detection of below threshold differences framed by contributions of variable clinical presentation to low reproducibility

**DOI:** 10.1101/2022.03.04.22271900

**Authors:** Anne K. Baker, Meghna Nanda, Su Hyoun Park, Katherine T. Martucci

**Affiliations:** Department of Anesthesiology, Human Affect and Pain Neuroscience Laboratory, Duke University School of Medicine; Center for Translational Pain Medicine, Duke Medical Center

**Author notes:** **Corresponding Author:** Katherine T. Martucci, PhD, Department of Anesthesiology, Duke Medical Center, Box DUMC 3094, Durham, NC 27710.

**Keywords:** MRI, Brain Structure, Chronic Pain, Medial Prefrontal Cortex, Anterior Cingulate Cortex

## Abstract

**Background:** Fibromyalgia is a prevalent chronic pain condition characterized by widespread pain and sensory hypersensitivity. While much remains unknown about fibromyalgia’s neurobiological underpinnings, central nervous system alterations appear to be heavily implicated in its pathophysiology. Previous research examining brain structural abnormalities associated with fibromyalgia has yielded inconsistent findings. Thus, we followed previous methods to examine brain gray matter differences in fibromyalgia. We hypothesized that, relative to healthy controls, participants with fibromyalgia would exhibit lower gray matter volume in regions consistently implicated in fibromyalgia: the anterior cingulate cortex and medial prefrontal cortex.

**Methods:** This study used magnetic resonance imaging to evaluate regional and whole brain differences in gray matter among females with and without fibromyalgia. Group differences were analyzed with two-sample t-tests, controlling for total intracranial volume.

**Results:** No significant differences in regional or whole brain gray matter volumes were detected between fibromyalgia and healthy controls.

**Conclusions:** Results add to an existing body of disparate findings regarding brain gray matter volume differences in fibromyalgia, and suggest structural differences previously detected in fibromyalgia should be examined for reproducibility. Absent significant differences may also suggest that functional, but not structural, brain adaptations are primarily associated with fibromyalgia.

## INTRODUCTION

Fibromyalgia (FM) is a common chronic pain condition.^1^ While the etiology of FM has yet to be fully elucidated, the presence of central sensitization appears to be a defining feature; this involves increased neuronal activity within the central nervous system (CNS) and exaggerated responses to incoming sensory input.^2^ In addition to functional CNS alterations, structural abnormalities may yield insight into CNS mechanisms relating to the development and maintenance of FM.

Neuroanatomical distinctions have been broadly observed in chronic pain conditions,^3,4^ including fibromyalgia.^5,6^ A recent meta-analysis of studies using voxel-based morphometry (VBM) to assess gray matter (GM) abnormalities associated with fibromyalgia reported significantly lower GM among FM patients relative to healthy controls in several brain regions, including 1) the bilateral anterior cingulate cortex (ACC) extending to the medial prefrontal cortex and paracingulate cortex, 2) the bilateral posterior cingulate cortex (PCC), and 3) the bilateral parahippocampal gyri extending to the fusiform cortex and hippocampus.^5^ Another seminal paper demonstrated similar results in the left ACC, right PCC, and right medial prefrontal cortex (mPFC), as well as in the bilateral thalamus and several other regions.^7^

While meta-analytic and ancillary findings largely report lower regional GM volumes associated with FM, others have reported greater GM in the striatum^8^ and left angular gyrus, right cuneus, and right postcentral gyrus.^7^ And still others have reported no GM differences when controlling for affective dysregulation.^9^ Baliki et al.^10^ have suggested it is improbable these discrepant findings are a function of differences among patients; rather, they point to variability in methods. However, given substantial variability in clinical presentations of FM,^11,12^ it is also plausible that varied FM symptomology may be associated with different underlying neurobiological mechanisms. Thus, in an effort to substantiate the reproducibility of previous results and lend insight into the question of patient versus methodological variability, the present study aimed to use VBM to closely follow previous methods^13,14^ and replicate previous findings of altered gray matter volumes in a sample of females with fibromyalgia (n=17) relative to sex-matched healthy control (HC) participants (n=17).

We hypothesized that, relative to HC participants, participants with FM would exhibit lower GM volume in regions consistently implicated in pain processing and, specifically, the ACC and the mPFC. Previous peak coordinates reported by Burgmer et al.^13^ and Ceko et al.^14^ are located within regions corresponding to the ACC and mPFC, but the precise locations are varied. Therefore, we first examined GMV in the whole brain. For the final manuscript, we will follow up on the whole brain analysis with region of interest (ROI) analysis focusing on the mPFC and ACC, using methods as in Baliki et al. (2011).

## METHODS

### Participants

A total of 35 participants completed the study procedures; however, data from one participant was excluded from analysis due to scan artifacts. Accordingly, data from 34 participants were included in the final analysis: 17 HC participants (mean age ± SD = 48.41 ± 10.3) and 17 with FM (mean age ± SD = 49.12 ± 9.6). All participants were female; sex was self-reported and not distinguished from gender.

Participants with FM were required to meet the following conditions of the 2011 American College of Rheumatology (ACR) criteria: (1) widespread pain index (WPI) >= 7 and symptom severity score (SSS) >= 5 or WPI between 3–6 and SSS >= 9; (2) symptoms present for at least 3 months; and (4) no other disorder explains that symptoms and pain being experienced. Additionally, FM participants had to have pain in all 4 quadrants of the body and an average pain score of 2 or greater over the previous month. FM participants were not taking any opioid medications, had not taken any opioid medications within 90 days prior to study visits, and had not taken any opioid medications for a period longer than 30 days in their lifetime. HC participants were required to have no chronic pain and no depression or anxiety. Other overall exclusion criteria were MRI contraindications, pregnancy or nursing, and claustrophobia.

### Study Procedures

All data collection and relevant study procedures, excluding data analysis, were completed at Stanford University in the Richard L. Lucas Center for Imaging. The Stanford Institutional Review Board approved all study procedures, and all participants were informed of the study details and signed written informed consent prior to initiating study procedures. A Data Use Agreement was established to share the data from Stanford University for analysis by the research team at Duke University.

Patients filled out the following questionnaires prior to MRI scanning: Beck Depression Inventory (BDI), State-Trait Anxiety Inventory (STAI State, STAI Trait), Behavioral Inhibition System/Behavioral Approach System (BIS/BAS Scales), Profile of Mood States (POMS), Positive and Negative Affect Schedule (PANAS), Fibromyalgia Assessment Form based on the Wolfe et al. Fibromyalgia Diagnostic Criteria, Brief Pain Inventory (BPI), and Patient-Reported Outcomes Measurement Information System (PROMIS, computer adapted test bank v1.0) Fatigue.

### Magnetic Resonance Imaging (MRI) Scans

A 3T General Electric MRI scanner with an 8-channel head coil (GE Systems, Chicago, IL) was used for acquiring a T1 anatomical scan (3D fast spoiled gradient-echo IRprep BRAVO). The anatomical scan sequence had the following parameters: whole brain coverage including the brainstem and cerebellum, 1 mm slice thickness, 22 mm frequency field of view, frequency direction anterior-posterior, number of excitations 2, flip angle 11°, TR 6.8, echo time 2.6, frequency 256, phase 256, and bandwidth 50.00.

### MRI Scan Preprocessing

Preprocessing was conducted using SPM12 according to the VBM Tutorial document provided by John Ashburner.^15^ Preprocessing was conducted in 3 steps: Segmentation, Running Dartel, and Normalization to MNI Space. All the modules were accessible via the SPM tab in the batching system.

Segmentation serves to identify the different types of tissues within the brain; SPM12 focuses on three main tissues: gray matter, white matter, and cerebral spinal fluid. The module was accessed by clicking on *SPM* → *Spatial* → *Segment*. All the HC and FM nifti files labelled “anat.nii” were selected in *Data* → *Channel* → *Volumes*. Under the *Tissues* section, the first three *Tissues* listed correspond to gray matter, white matter, and CSF respectively. *Native Tissue* was changed to *Native + Dartel imported* for the gray matter and white matter *Tissue* but was changed to *Native Space* for the CSF *Tissue*. The following settings were left at default: (1) *Bias regularization, BIAS FWHM, Save Bias Corrected* for the *Channel* section; (2) all settings for *Tissue* that corresponded to skull, soft tissue outside brain, and extra outside the head; (3) *MRF Parameter, Clean Up Warping regularization, Affine regularization, Sampling distance*, and *Deformation fields* under the *Warping & MRF* section. The Segmentation module returned c1, c2, and c3 nifti files that correspond to the gray matter, white matter, and CSF tissues respectively. The module also returned rc1, rc2, and rc3 nifti files that were used for the later Run Dartel step.

The Run Dartel step creates its own average template and aligns the data from the rc1 and rc2 files, therefore aligning the gray and white matter. These files were selected under the *Images* section. All the values under the *Settings* section were not changed and were left at default. The Run Dartel module returned u_rc1 files as well as 6 template files.

The file named “Template 6” was used for the MNI Normalization step under the Normalize to MNI Space module under *Dartel Template*. Only the u_rc1 files were chosen under *Many Subjects* → *Flow fields* and c1 files for *Images*. Additional setting changes included selecting *Preserving Amount* for the *Preserve* section and using a Gaussian size of 8mm under the *Gaussian FWHM* section. All other settings such as *Voxel Sizes* and *Bounding box* were left at default. This module returned smwc1 files that would be used for the final analysis.

### Power Analysis

Prior to data analysis and included in the pre-registration plan (https://archive.org/details/osf-registrations-phet2-v1, 10.17605/OSF.IO/PHET2), a power analysis was conducted using data obtained from Burgmer et al. (2009). Power analysis indicated that with an alpha less than 0.05, power of at least 0.8, and both group sizes as n = 17, this analysis was sufficiently powered to detect a medium effect size with r^2^ = 0.4.

### MRI Scan Between-Group Analysis

To obtain differences in brain gray matter volume (GMV) between HC and participants with FM, we performed a two-sample t-test using the factorial design specification interface in SPM12 running on Matlab R2020b. FM data were entered as group 1, HC data were entered as group 2, and total intracranial volume was entered as a covariate of no interest. Consistent with the steps outlined in the VBM manual, we used absolute masking and a threshold of 0.01.

## RESULTS

### General Characteristics

General characteristics of patients with FM and HC participants are shown in **Table 1**. Age and differences in emotional states were evaluated using independent samples t-tests. There was a significant difference in mean PANAS negative affect scores (t_(1,16)_ = 3.70, *p* = 0.002), with patients with FM reporting higher levels of negative affect than HC participants. Similarly, patients reported significantly lower levels of positive affect than HC participants (t_(1,16)_ = -3.75, *p* = 0.002). We also observed significant differences in POMS-TMD scores (t_(1,16)_ = 5.89, *p* < 0.01) and in BDI scores (t_(1,16)_ = 5.62, *p* < 0.01), with patients reporting significantly greater mood disturbance and depressive symptoms. PROMIS Fatigue scores were also significantly higher among patients relative to HC participants (t_(1,17)_ = 6.04, *p* < 0.001). Finally, as expected, patients exhibited significantly greater BPI Pain Severity (t_(1,16)_ = 9.27, *p* < 0.01) and significantly greater BPI Pain Interference (t_(1,16)_ = 6.97, *p* < 0.01).

**Table 1.**
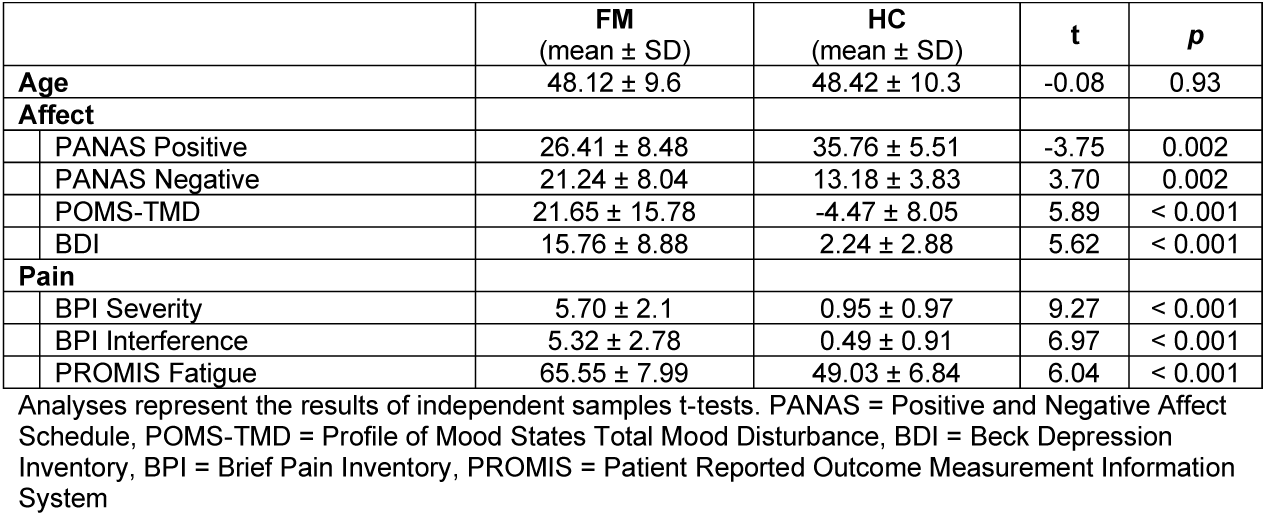
Participant Characteristics

### Voxel-based Morphometry Analysis

An independent samples t-test controlling for total intracranial volume revealed no significant GM differences between FM patients and HC participants. However, one cluster (x = 54, y = -9, z = 3) approached significance at *p* = 0.057 (Figure 1). Additional non-significant suprathreshold clusters are reported in **Table 2**.

**Figure 1.**
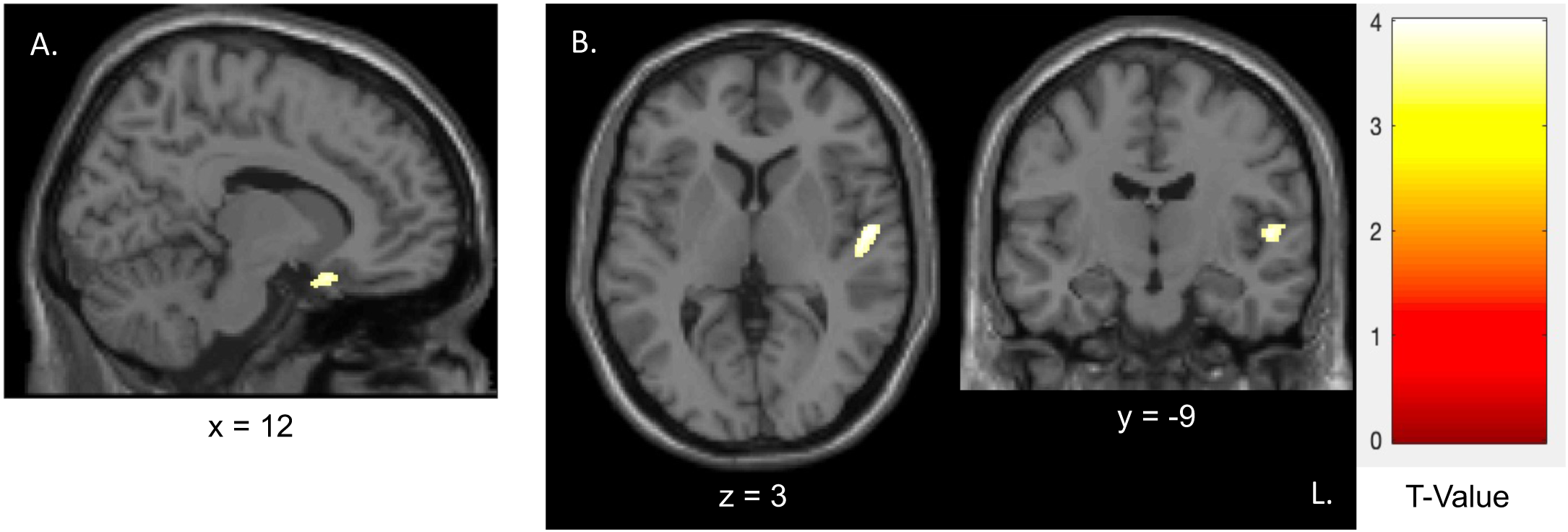
Non-significant GM Differences Detected by VBM. **Whole-brain VBM analysis identified non-significant clusters of interest in regions relevant to pain. A**. GM differences in frontal orbital cortex, within perigenual ACC, showing GM in FM < HC, *p* = 0.31 (uncorrected), extent threshold *K* = 80. **B**. GM differences approaching significance in the transverse temporal gyrus when FM < HC, *p* = 0.057 (uncorrected), extent threshold *K* = 309. See also coordinates and peak voxel statistics in Table 2. Abbreviations: ACC, anterior cingulate cortex; GM, gray matter; HC, healthy controls; FM, fibromyalgia; VBM, voxel-based morphometry; T-Value significance threshold uncorrected.

**Table 2.** Whole Brain Suprathreshold Clusters, Uncorrected

| Brain region | MNI coordinates (x, y, z) |  |  | Cluster size (voxels) | p value of cluster | Peak t value |
| --- | --- | --- | --- | --- | --- | --- |
| Supramarginal Gyrus | -52 | -46 | 39 | 216 | 0.105 | 4.43 |
| Precentral Gyrus | -33 | -4 | 72 | 106 | 0.245 | 4.39 |
| White Matter | 28 | -16 | 24 | 147 | 0.174 | 4.35 |
| Transverse Temporal Gyrus | 54 | -9 | 3 | 309 | 0.057 | 4.15 |
| Cerebellum | -10 | -50 | -28 | 45 | 0.452 | 4.00 |
| Frontal Orbital Cortex | 12 | 15 | -21 | 80 | 0.312 | 3.74 |
| Frontal Orbital Cortex | 34 | 27 | -21 | 120 | 0.217 | 3.73 |

Brain regions were identified using the Harvard-Oxford cortical structural atlas based on peak MNI coordinates. All clusters had peak level p-values, uncorrected, <0.001. Clusters with <45 voxels were not included.

## DISCUSSION

The goal of the present study was to replicate previous observations that FM patients exhibit lower gray matter volumes in specific brain regions, namely the ACC and mPFC. We conducted a power analysis and included a rigorous sample of high-quality structural MRI data sets in patients with FM vs. pain-free HC participants. Despite adequate power and a sample size comparable to previous VBM studies of patients with fibromyalgia, we were unable to replicate published findings of altered brain GM structure in FM. Given that it is important to publish non-significant findings, we report our null findings and discuss the potential contribution of individual differences—a particularly relevant confound in fibromyalgia—to variable success in identifying reliable and reproducible GM structural alterations.

### Overview of Fibromyalgia and Variability Inherent in the Clinical Population

Fibromyalgia is a chronic pain condition characterized by widespread, generalized musculoskeletal pain. The overarching diagnostic criteria for FM, as defined for clinical research and approved by the ACR, are generalized pain in at least 4 of 5 regions of the body (4 quadrants plus axial pain), symptom duration of at least 3 months, and either a widespread pain index (WPI) score ≥ 7 combined with a symptom severity scale (SS) score ≥ 5, or a WPI score ranging from 4 to 6 combined with a SSS score ≥ 9.^16^ Individuals meeting these three criteria, with no other condition that would fully explain their symptoms, meet standardized research criteria for FM regardless of comorbid illnesses. While these broad criteria provide a useful framework for categorizing clinical presentations for research studies, they do not fully capture the various combinations and permutations of FM symptoms relating to pain itself nor the many common non-painful symptoms associated with FM such as fatigue, cognitive dysfunction, emotional disturbances, and sleep impairment. For example, while the severity and some aspects of pain and other symptoms are captured in the WPI and SS scores of the ACR FM research criteria, the relative contributions of these symptoms and effects additional comorbid symptoms and conditions are not captured. Meanwhile, to date, individual research studies have not yet consistently used the ACR FM research criteria (described above) to identify eligible participants with FM. Indeed many studies rely on physician diagnosis of FM as inclusion criteria, which may be susceptible to individual physician views and opinions. Accurate identification of the neurobiological antecedents and correlates of FM, as well as the identification of potential uniquely phenotyped subgroups of FM, may hinge on this variability.

Similarly, both genomics and environmental factors have been implicated in the pathophysiology of FM. It is not yet clear whether one causal factor alone is a necessary and sufficient condition for development of FM or if gene-environment interactions generate epigenetic alterations that in turn facilitate experiences of FM. Buskila et al.^17^ report high familial aggregation of FM, positing polygenic risk factors that include serotonergic, dopaminergic and catecholaminergic systems. However, while specific gene candidates associated with FM have indeed been identified^18^ and include potentially important DNA patterns (e.g., hypomethylation),^19^ there is simultaneous acknowledgment of strong evidence pointing to environmental factors. For example, recent meta-analytic findings show a significant association between FM and exposure to traumatic stress,^20^ and a systematic review reported that psychological trauma frequently appears to immediately precede onset of FM.^21^ These potentially distinctive causal pathways may differentially alter brain structure and function, yielding an assortment of clinical presentations that are relatively unalike despite fitting the FM diagnostic criteria for research.

### Central Nervous System Changes Observed in Patients with Fibromyalgia

Fibromyalgia has been broadly associated with CNS changes, including abnormalities in central pain processing (e.g., central sensitization) that commonly manifest as hyperalgesia and allodynia.^22–24^ Structural changes in brain regions heavily implicated in pain and sensory processing may be related to the development of central sensitization and its experiential sequelae. More specifically, both meta-analytic and focused study findings using VBM, volumetrics and/or multi-modal neuroimaging approaches show FM-related reduced GMV in regional hubs of both ascending and descending pain pathways such as the insula, ACC, PCC, parahippocampal gyrus, and mPFC.^5,7,25^ While reduced GMV in the ACC and mPFC appear to be somewhat consistent, the demographic and clinical profiles reported in previous studies vary considerably and may impact the locations of significant findings.

For example, meta-analytic results showing GM differences in the medial pain system and default mode network reported by Shi et al.^5^ incorporated seven studies with samples that included FM patients with and without affective disorders, patients with an average age less than 50 years old, and patients with an average age greater than 50 years old. Pomares et al.^7^ reported significant GM differences in several cortical regions, including the ACC and mPFC, between FM patients and pain-free controls, however their sample consisted only of post-menopausal females, thereby removing the possible impact of fluctuating sex hormones on pain and related brain structure and function.^26–28^ Moreover, consistent with previous research, Pomares et al.^7^ reported significant differences in Beck Depression Inventory scores and Pain Catastrophizing scores, among others. Given that Hsu et al.^9^ have indicated the absence of consistent differences in GMV between FM patients and healthy controls when controlling for affective disorders, it may be that affect-related neuroadaptations subserved GM differences in FM observed by Pomares et al.—who included only age as a covariate—as well as by several other VBM studies that examined but did not covary psychological factors.

Based on these differences in methods and covariates included in published research of GM differences in FM, it is clear that future studies should examine the extent to which psychological factors, age, sex hormones, and other consistently reported variables impact VBM analyses, ideally in large meta-analytic studies. Such research may provide important and clear connections between the myriad factors related to FM and GM structural changes, as well as provide insight into subgroups of FM based on the presence/absence of GM structural differences and covarying factors.

### Potential Contributions of Patient Variability to a Lack of Significant VBM Group Results

In addition to the use of VBM structural analyses to aid in understanding chronic pain conditions such as FM, the usefulness of VBM has been demonstrated for other clinical conditions including Alzheimer’s Disease and schizophrenia.^29^ Such conditions may impart more observable and/or more consistent brain changes compared to patients with FM. Furthermore, these other conditions may be more consistent across patients, perhaps with less variety of clinical and symptom presentations than typically found in FM cohorts. All to say, it is possible that variability of pain locations, durations, additional psychological factors and a variety of medication use may be contributing to non-significant GM differences observed in our study.

### Importance of Reporting Non-Significant Findings

Although our results did not support our initial hypotheses, reporting these results is prudent, particularly to allow for more balanced literature on the presence or absence of GM differences detected by VBM in patients with FM. Indeed, a trend for pre-registration of analyses has led to a greater number of reported non-significant findings.^30^ A recent preprint concluded that 61% of surveyed studies that pre-registered their analyses reported results that did not support their hypotheses^31^—a rate much higher than previously estimated rates or reported results that did not support the initial hypothesis.^32,33^

### Limitations

A few limitations should be noted in consideration of these results. While our power analysis indicated a sufficient sample size, it may be that the variability of patient chronic pain conditions described above (i.e., pain locations, comorbid psychological and clinical conditions, medication use) requires a larger sample size to identify regional GM differences in FM. A larger sample would also afford additional statistical power for including additional covariates in analyses. Additionally, we were unable to account for the role of sex hormones and variability across the menstrual cycle. Menstrual cycle tracking and/or blood collection for hormone monitoring will be an essential aspect of future studies to minimize the potential influence of menstrual cycle variability on study results.^26–28^ Finally, patients were taking a variety of non-opioid medications, including serotonin-norepinephrine reuptake inhibitors (SNRIs; e.g., duloxetine, venlafaxine) and GABA analogues (e.g., gabapentin, pregabalin). Previous research suggests these medications are associated with changes in brain structure, including reduced gray matter volume associated with pregabalin administration among FM patients,^34^ and altered gray matter associated with duloxetine administration in osteoarthritis^35^ and chronic low pain back pain.^36^ Accordingly, medication use may also be a critical confound in VBM and should examined among larger samples in the future.

## Conclusions

Despite limitations inherent to clinical neuroimaging research, the present study contributes valuable null findings to the existing body of literature exploring GM differences in FM using VBM. Nested within a framework of previous research with mixed findings, non-significant whole-brain results suggest there may be important individual differences and/or distinctive methodological approaches that impact detection of significant differences in FM with VBM.

## Data Availability

All data produced in the present work are contained in the manuscript.

## Acknowledgements

We thank Daniel Weikel, MS, Duke University Department of Anesthesiology, Biostatistics Core for his assistance with calculating the power analysis, and Cristina (Cojocaru) Mackey, DO, and Erin Perrine, BS, for their assistance with data collection. Special thanks to Sean Mackey, MD, PhD, at Stanford University, for additional support of the study and to Kevin Johnson, RN, PhD, at Florida State University, for his advice regarding the VBM analysis. We especially thank all of the participants for their generosity of time and efforts spent in the study.

## REFERENCES

1. Walitt B, Nahin RL, Katz RS, Bergman MJ, Wolfe F. The Prevalence and Characteristics of Fibromyalgia in the 2012 National Health Interview Survey. PLoS One. 2015;10(9):e0138024. doi:10.1371/journal.pone.0138024

2. Latremoliere A, Woolf CJ. Central sensitization: a generator of pain hypersensitivity by central neural plasticity. J Pain. 2009;10(9):895–926. doi:10.1016/j.jpain.2009.06.012

3. Apkarian AV, Sosa Y, Sonty S, et al. Chronic back pain is associated with decreased prefrontal and thalamic gray matter density. J Neurosci. 2004;24(46):10410–10415. doi:10.1523/JNEUROSCI.2541-04.2004

4. Kim JH, Suh SI, Seol HY, et al. Regional grey matter changes in patients with migraine: a voxel-based morphometry study. Cephalalgia. 2008;28(6):598–604. doi:10.1111/j.1468-2982.2008.01550.x

5. Shi H, Yuan C, Dai Z, Ma H, Sheng L. Gray matter abnormalities associated with fibromyalgia: A meta-analysis of voxel-based morphometric studies. Semin Arthritis Rheum. 2016;46(3):330–337. doi:10.1016/j.semarthrit.2016.06.002

6. McCrae CS, O’Shea AM, Boissoneault J, et al. Fibromyalgia patients have reduced hippocampal volume compared with healthy controls. J Pain Res. 2015;8:47–52. doi:10.2147/JPR.S71959

7. Pomares FB, Funck T, Feier NA, et al. Histological Underpinnings of Grey Matter Changes in Fibromyalgia Investigated Using Multimodal Brain Imaging. J Neurosci. 2017;37(5):1090–1101. doi:10.1523/JNEUROSCI.2619-16.2016

8. Schmidt-Wilcke T, Luerding R, Weigand T, et al. Striatal grey matter increase in patients suffering from fibromyalgia--a voxel-based morphometry study. Pain. 2007;132 Suppl 1:S109–S116. doi:10.1016/j.pain.2007.05.010

9. Hsu MC, Harris RE, Sundgren PC, et al. No consistent difference in gray matter volume between individuals with fibromyalgia and age-matched healthy subjects when controlling for affective disorder. Pain. 2009;143(3):262–267. doi:10.1016/j.pain.2009.03.017

10. Baliki MN, Schnitzer TJ, Bauer WR, Apkarian AV. Brain morphological signatures for chronic pain. PLoS One. 2011;6(10):e26010. doi:10.1371/journal.pone.0026010

11. Bartley EJ, Robinson ME, Staud R. Pain and Fatigue Variability Patterns Distinguish Subgroups of Fibromyalgia Patients. J Pain. 2018;19(4):372–381. doi:10.1016/j.jpain.2017.11.014

12. Toussaint L, Vincent A, McAllister SJ, Whipple M. Intra- and Inter-Patient Symptom Variability in Fibromyalgia: Results of a 90-Day Assessment. Musculoskeletal Care. 2015;13(2):93–100. doi:10.1002/msc.1090

13. Burgmer M, Gaubitz M, Konrad C, et al. Decreased gray matter volumes in the cingulo-frontal cortex and the amygdala in patients with fibromyalgia. Psychosom Med. 2009;71(5):566–573. doi:10.1097/PSY.0b013e3181a32da0

14. Ceko M, Bushnell MC, Fitzcharles MA, Schweinhardt P. Fibromyalgia interacts with age to change the brain. Neuroimage Clin. 2013;3:249–260. doi:10.1016/j.nicl.2013.08.015

15. VBMclass15.pdf. https://www.fil.ion.ucl.ac.uk/~john/misc/VBMclass15.pdf

16. Wolfe F, Clauw DJ, Fitzcharles MA, et al. 2016 Revisions to the 2010/2011 fibromyalgia diagnostic criteria. Semin Arthritis Rheum. 2016;46(3):319–329. doi:10.1016/j.semarthrit.2016.08.012

17. Buskila D, Sarzi-Puttini P. Biology and therapy of fibromyalgia. Genetic aspects of fibromyalgia syndrome. Arthritis Res Ther. 2006;8(5):218. doi:10.1186/ar2005

18. D’Agnelli S, Arendt-Nielsen L, Gerra MC, et al. Fibromyalgia: Genetics and epigenetics insights may provide the basis for the development of diagnostic biomarkers. Mol Pain. 2019;15:1744806918819944. doi:10.1177/1744806918819944

19. Ciampi de Andrade D, Maschietto M, Galhardoni R, et al. Epigenetics insights into chronic pain: DNA hypomethylation in fibromyalgia-a controlled pilot-study. Pain. 2017;158(8):1473–1480. doi:10.1097/j.pain.0000000000000932

20. Kaleycheva N, Cullen AE, Evans R, Harris T, Nicholson T, Chalder T. The role of lifetime stressors in adult fibromyalgia: systematic review and meta-analysis of case-control studies. Psychol Med. 2021;51(2):177–193. doi:10.1017/S0033291720004547

21. Yavne Y, Amital D, Watad A, Tiosano S, Amital H. A systematic review of precipitating physical and psychological traumatic events in the development of fibromyalgia. Semin Arthritis Rheum. 2018;48(1):121–133. doi:10.1016/j.semarthrit.2017.12.011

22. Staud R, Spaeth M. Psychophysical and neurochemical abnormalities of pain processing in fibromyalgia. CNS Spectr. 2008;13(3 Suppl 5):12-17. doi:10.1017/s109285290002678x

23. Harte SE, Harris RE, Clauw DJ. The neurobiology of central sensitization. J Appl Biobehav Res. 2018;23(2):e12137. doi:10.1111/jabr.12137

24. Hazra S, Venkataraman S, Handa G, et al. A Cross-Sectional Study on Central Sensitization and Autonomic Changes in Fibromyalgia. Front Neurosci. 2020;14:788. doi:10.3389/fnins.2020.00788

25. Robinson ME, Craggs JG, Price DD, Perlstein WM, Staud R. Gray matter volumes of pain-related brain areas are decreased in fibromyalgia syndrome. J Pain. 2011;12(4):436–443. doi:10.1016/j.jpain.2010.10.003

26. Xiao X, Yang Y, Zhang Y, Zhang XM, Zhao ZQ, Zhang YQ. Estrogen in the anterior cingulate cortex contributes to pain-related aversion. Cereb Cortex. 2013;23(9):2190–2203. doi:10.1093/cercor/bhs201

27. Straube T, Schmidt S, Weiss T, Mentzel HJ, Miltner WHR. Sex differences in brain activation to anticipated and experienced pain in the medial prefrontal cortex. Hum Brain Mapp. 2009;30(2):689–698. doi:10.1002/hbm.20536

28. Tu CH, Niddam DM, Chao HT, et al. Brain morphological changes associated with cyclic menstrual pain. Pain. 2010;150(3):462–468. doi:10.1016/j.pain.2010.05.026

29. Scarpazza C, De Simone MS. Voxel-based morphometry: current perspectives. Nano. 2016;5:19–35. doi:10.2147/NAN.S66439

30. Mehler DMA, Edelsbrunner PA, Matić K. Appreciating the significance of non-significant findings in psychology. J Eur Psychol Stud. 2019;10(4):1. doi:10.5334/e2019a

31. Allen C, Mehler DMA. Correction: Open science challenges, benefits and tips in early career and beyond. PLoS Biol. 2019;17(12):e3000587. doi:10.1371/journal.pbio.3000587

32. Cristea IA, Ioannidis JPA. P values in display items are ubiquitous and almost invariably significant: A survey of top science journals. PLoS One. 2018;13(5):e0197440. doi:10.1371/journal.pone.0197440

33. Fanelli D. Negative results are disappearing from most disciplines and countries. Scientometrics. 2012;90(3):891–904. doi:10.1007/s11192-011-0494-7

34. Puiu T, Kairys AE, Pauer L, et al. Association of Alterations in Gray Matter Volume With Reduced Evoked-Pain Connectivity Following Short-Term Administration of Pregabalin in Patients With Fibromyalgia. Arthritis Rheumatol. 2016;68(6):1511–1521. doi:10.1002/art.39600

35. Tetreault P, Baliki M, Vachon-Presseau E, Yeasted R, Schnitzer TJ, Apkarian A. Cortical singularity of knee osteoarthritis patients and their response to duloxetine or placebo treatment. Osteoarthritis Cartilage. 2015;23:A354. doi:10.1016/j.joca.2015.02.653

36. Chatterjee N, Noor N, Crowell A, Mackey S. Duloxetine and placebo alter different gray matter regions in chronic low back pain patients. J Pain. 2010;11(4):S40. doi:10.1016/j.jpain.2010.01.169

